# Bivalent mRNA vaccine effectiveness against COVID-19 infections, hospitalisations and deaths in Portugal: a cohort study based on electronic health records, September 2022 to May 2023

**DOI:** 10.1101/2023.09.05.23295025

**Authors:** Ausenda Machado, Irina Kislaya, Patricia Soares, Sarah Magalhães, Ana Paula Rodrigues, Rafael Franco, Pedro Pinto Leite, Carlos Matias Dias, Baltazar Nunes

## Abstract

In Portugal, a bivalent COVID-19 vaccine booster was recommended for those with complete primary COVID-19 vaccination, starting on September 6 2022. This study aims to estimate the mRNA bivalent vaccine effectiveness (VE) against COVID-19 infection, hospitalisation and death in the Portuguese population aged 65 and more years with a follow-up of more than six months.

**Methods:** We used a cohort approach to analyse six electronic health registries using deterministic linkage. The follow-up period comprehend September 2022 to May 2023. The outcomes included SARS-CoV-2 infection, COVID-19-related hospitalisation and death. Individuals were considered vaccinated 14 days following a bivalent mRNA COVID-19 vaccine uptake. For each outcome, COVID-19 bivalent VE was estimated as one minus the confounder adjusted hazard ratio of bivalent vaccine vs no bivalent vaccine, estimated by Cox regression with time-dependent vaccine exposure.

**Results:** In the ≥ 80 year-olds, bivalent VE was 23.2 (95%CI: 20.1 to 26.2), 41.3 (95%CI: 34.5 to 47.5) and 50.3 (44.6 to 55.3), against infection, COVID-19-related hospitalisation and death, respectively. In the 65-79 year-old, bivalent VE against infection was 37.7 (35.5 to 39.8), 58.5 (95%CI: 51.9 to 64.2) against hospitalisation and 65.1 (95%CI: 59 to 70.4) against death. Vaccine effectiveness decay was observed for both age groups and in all outcomes, up to 6 months of vaccine uptake.

**Conclusions:** In a population with a high risk of SARS-CoV-2 complications, we observed moderate bivalent VE estimates against severe COVID-19 and low protection against infection. The lower VE estimates observed in the ≥ 80 year-olds should be interpreted in light of the reference group used for the estimation, i.e., individuals with high vaccine coverage (both primary series and multiple boosters). Significant VE decay was observed up to six months of vaccine uptake, which should be considered when preparing future vaccination campaigns.

## Introduction

On September 6 2022, Portuguese health authorities recommended COVID-19 booster vaccination with adapted bivalent mRNA vaccines to those aged 60 or more (1) to match the circulation virus variant and thus prevent severe complications after Omicron SARS-CoV-2 infection. Besides age criteria, individuals with complete primary vaccination were eligible for the seasonal booster regardless of the previous history of boosters uptake but provided that a minimum interval of 3 months from a previous SARS-COV-2-related event (infection or vaccination) was completed. The first available bivalent vaccine was from Comirnaty Original/BA.1 (2), followed by Comirnaty Original/ BA.4-5 (3) and Spikevax Original/BA.1 (4). The last approved vaccine was Spikevax Original/ BA.4-5 (in late November 2022). Vaccination campaign with bivalent vaccines started on September 7, 2022 and, as previously, was rolled out by age criteria, prioritising those aged 80 and more years.

Considering the previous monovalent COVID-19 vaccination in Portugal, the primary series vaccination started in February 2021 and achieved extremely high vaccine coverage, with more than 90% of the population aged 65 and more considered with complete vaccination. Throughout the pandemic, different booster doses were recommended to this group of the population, but the last booster campaign (sprint booster in May 2022) was only recommended to the 80 and more years (5) (reaching a 68% coverage by week 36 2022 (6)).

In September 2022, the epidemiological situation in Portugal was characterised by the predominant circulation of the Omicron BA.5 variant. During the autumn and winter months, the BA.5 variant decreased frequency and XXQ surged in week 40/2022. By week 10/2023 more than 50% of the sequenced virus were classified as Omicron-XBB variant (7). XBB variant has been described as a potential increase of immunity evasion (8).

In the context of the introduction of new vaccines, adapted bivalent, different variants in circulation, and the preparation of future vaccination campaigns, it is important to evaluate the protection against severe outcomes granted by the vaccine used in the national programme, particularly in the population with increased risk of complications.

This study aims to estimate vaccine effectiveness (VE) of the adapted bivalent mRNA vaccines against infection, COVID-19 hospitalisation and COVID-19-related death for the Portuguese population with more than 65 years old and by time since vaccination between September 7 2022 and May 31 2023.

## Methods

### Study Design

A cohort approach was used to estimate VE of bivalent mRNA booster, using electronic health registries. The study design was described previously in previous studies (9–11), but in summary, a deterministic data linkage of electronic health records was done using the unique national health number.

The target population included those residents in mainland Portugal, aged 65-110 years, who completed the primary vaccination scheme and were eligible for bivalent booster vaccination. The following exclusion criteria were adopted: being non-users of the National Health System (NHS), being infected with SARS-COV-2 within 90 days before the start of the follow-up, being vaccinated with monovalent vaccines within 90 days before the start of the seasonal vaccination campaign, being vaccinated with the monovalent vaccines during the follow-up period, and having incomplete or inconsistent data on vaccination (interval between 2 doses for primary vaccination less than 19 days, brands not recommended in Portugal, interval between successive booster vaccinations less than 90 days, a combination of brands other than recommended, vaccinated with bivalent vaccines before authorised in Portugal, partially vaccinated (with one dose only of 2-dose vaccine)) (Figure flowchart).

The follow-up period was between September 7 2022 and May 31 2023, defined based on the vaccination campaign calendar. The data extraction and linkage for this study was performed by the Serviços Partilhados do Ministerio da Saude (SPMS) on July 27 2023, to allow data consolidation.

### Outcome and Exposure definitions

Three outcomes of interest were considered: 1) a laboratory-confirmed infection with SARS-CoV-2 notified to the National Surveillance System (SINAVE) regardless of the presence of symptoms; 2) COVID-19 hospitalisation, defined as an admission to a hospital for at least 24 hours, following laboratory-confirmed infection with SARS-CoV-2 and having COVID-19 as the primary diagnosis at discharge (ICD10 coding U071); and 3) COVID-19-related death, defined as death for which COVID-19 was recorded as the cause of death (U071) or deaths that occurred within 30 days after the laboratory-confirmed SARS-CoV-2 infection.

The exposure was an uptake of bivalent mRNA vaccine (Pfz/BA1, Pfz/BA5, Mod/BA1, Mod/BA5). Individuals were considered vaccinated 14 days following a bivalent mRNA vaccine uptake. Time since a bivalent vaccine was considered to evaluate the hypothesis of VE waning, VE was estimated for 14-97 days, 98-181 days and ≥180 days after bivalent vaccination.

Participants eligible for but without bivalent booster uptake (regardless of their history of previous boosters) were considered as a reference group. Further this group is referred to as “without bivalent vaccine”.

### Statistical analysis

Descriptive statistics were used to characterise participants at baseline by exposure level and at the end of the follow-up period. We performed a Cox regression adjusted for sex, age group, region, municipality deprivation index quintile, history of influenza and pneumococcus vaccination in the previous three years, number of COVID-19 tests in 2020-2022, comorbidities (without, immunocompromised, not immunocompromised), previous infection (previous positive test before the start of the follow up). Time period was included as strata in the model, considering intervals of 14 days. VE was estimated as one minus the confounder adjusted hazard ratio of bivalent vaccine vs no bivalent vaccine. Complete case analysis was used. Data analysis was performed with R software, version 4.0.5 (R Foundation, Vienna, Austria). The statistical significance level was set at 5%.

### Ethical statement

The study received approval from the Ethical Committee and the Data Protection Officer of the Instituto Nacional de Saúde Doutor Ricardo Jorge (December 13 2022). All data were anonymised by the SPMS before made available for statistical analysis at INSA.

## Results

We included in the analysis 2,151,531 individuals aged 65 and more years, 513,149 unvaccinated and 1,638,382 vaccinated. Figure 1 presents how many individuals were excluded for each exclusion criteria. The vaccine roll out indicate that maximum vaccine uptake plateau was in mid-October for the 80 and more years (1 month after vaccination started) and mid-December for the 65-79 years (4 month after vaccination started). Of the vaccinated individuals, 52.9% had the Comirnaty Origin/BA.1, 42.8% the Comirnaty Origin/BA 4-5 and 4.3% the Spikevax Origin/BA.1. Their distribution along time are in accordance with the availability and respective disposal of each vaccine brand/ composition.

**Figure 1:**
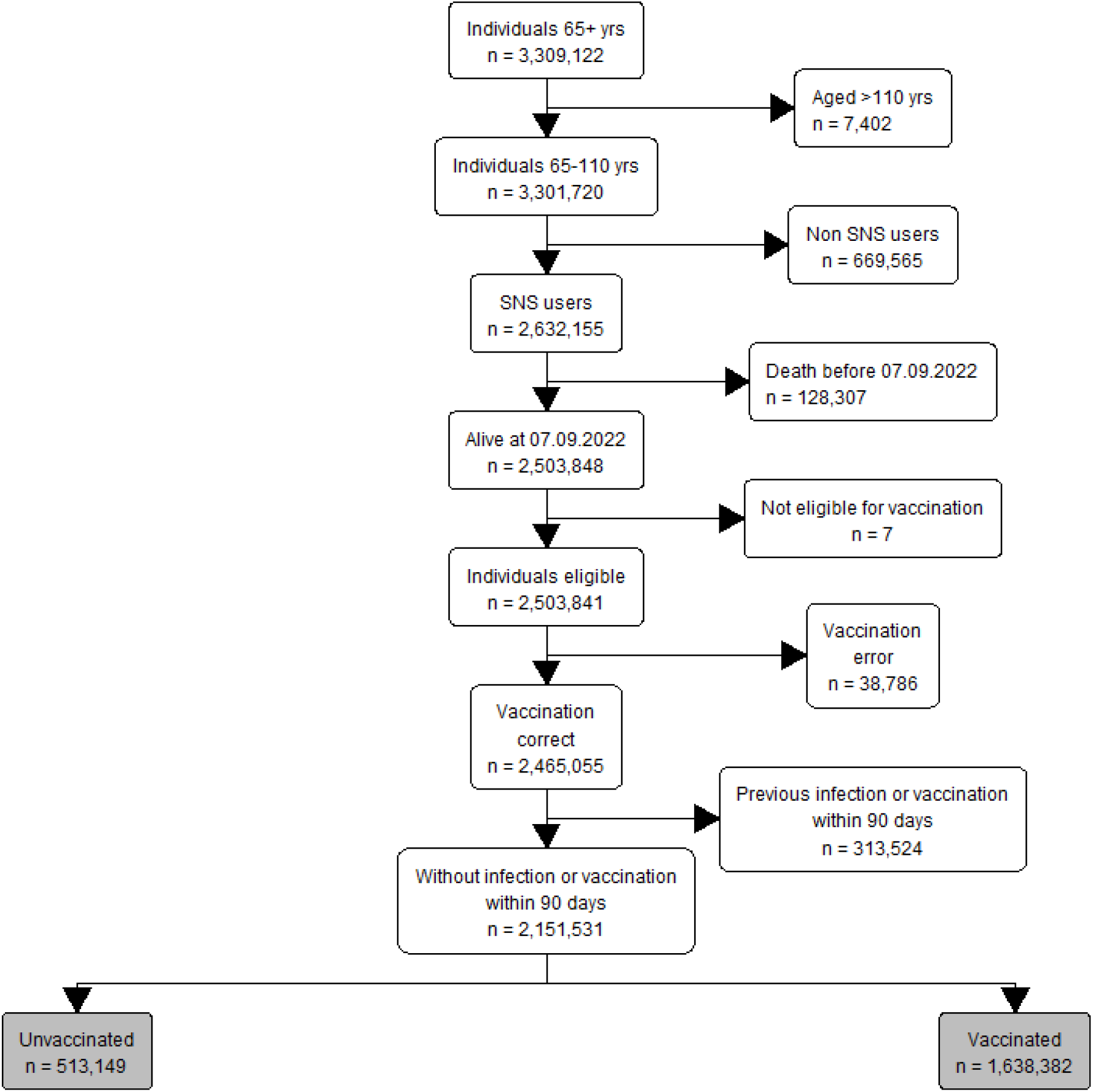
Flowchart of the study indicating inclusion/exclusion criteria.

The comparison of individuals with and without bivalent vaccines reveals a higher proportion of comorbidities and older age among the first group (Table 1).

**Table 1.** Description of the population during the study period – September 7 2022 to March 31 2023.

|  | Without bivalent vaccine | With bivalent booster |
| --- | --- | --- |
|  | No. 513,149 | No. 1,638,382 |
| Age (years), median (IQR) | 73.0 (68.0 - 79.0) | 74.0 (69.0 - 81.0) |
| **Age group, (%)** |  |  |
| 65-69 | 169,026 (32.9%) | 411,519 (25.1%) |
| 70-74 | 124,841 (24.3%) | 414,041 (25.3%) |
| 75-79 | 94,596 (18.4%) | 338,853 (20.7%) |
| 80-84 | 62,358 (12.2%) | 230,856 (14.1%) |
| 85-89 | 41,460 (8.1%) | 156,310 (9.5%) |
| 90-94 | 16,643 (3.2%) | 68,077 (4.2%) |
| 95+ | 4,225 (0.8%) | 18,726 (1.1%) |
| Sex Male, n (%) | 198,108 (38.6%) | 732,171 (44.7%) |
| **Region, n (%)** |  |  |
| A.R.S. Alentejo | 31,100 (6.1%) | 84,887 (5.2%) |
| A.R.S. Algarve | 38,977 (7.6%) | 60,447 (3.7%) |
| A.R.S. Centro | 99,911 (19.5%) | 313,052 (19.1%) |
| A.R.S. LVT | 175,873 (34.3%) | 571,935 (34.9%) |
| A.R.S. Norte | 161,128 (31.4%) | 600,853 (36.7%) |
| Missing | 6,160 (1.2%) | 7,208 (0.4%) |
| **EDI Quintile, n (%)** |  |  |
| Q1 (least deprived) | 74,745 (14.6%) | 255,425 (15.6%) |
| Q2 | 77,970 (15.2%) | 245,686 (15.0%) |
| Q3 | 76,846 (15.0%) | 241,289 (14.7%) |
| Q4 | 134,906 (26.3%) | 474,169 (28.9%) |
| Q5 (most deprived) | 142,522 (27.8%) | 414,605 (25.3%) |
| Missing | 6,160 (1.2%) | 7,208 (0.4%) |
| **Comorbidity, n (%)** |  |  |
| no | 126,964 (24.7%) | 304,773 (18.6%) |
| lowR | 279,885 (54.5%) | 908,963 (55.5%) |
| highR | 106,300 (20.7%) | 424,646 (25.9%) |
| **Number of SARS-CoV-2 tests, n (%)** |  |  |
| 0 | 200,685 (39.1%) | 518,258 (31.6%) |
| 1 | 109,869 (21.4%) | 346,270 (21.1%) |
| 2 | 68,827 (13.4%) | 237,301 (14.5%) |
| 3 | 42,279 (8.2%) | 156,393 (9.5%) |
| 4-9 | 73,734 (14.4%) | 300,454 (18.3%) |
| 10+ | 17,755 (3.5%) | 79,706 (4.9%) |
| Any other vaccine, n (%)<sup>†</sup> | 288,735 (56.3%) | 1,493,959 (91.2%) |
| Previous infection, n (%) | 95,070 (18.5%) | 436,656 (26.6%) |
| **Vaccine type, n (%)** |  |  |
| Spikevax Origin/BA.1 | NA | 70,440 (4.3%) |
| Comirnaty Origin/BA.1 | NA | 867,415 (52.9%) |
| Comirnaty Origin/BA 4-5 | NA | 700,527 (42.8%) |

<sup>†</sup> received at least one of the following vaccines since 2019: influenza, PN23, PCV7, 10 or 13
NA- not applicable

A total of 45,933 SARS-CoV-2 infections (65% in the 65-79 years), 2,865 COVID-19 hospitalisations (36.7% in the 65-79 years) and 2,615 deaths (30.2% in the 65-79 years) were registered during the follow-up. Moreover, the decline in hospitalisation may also be related to delays updating the hospital discharge database.

For the period in analysis, bivalent VE was estimated as 23.2 (95%CI: 20.1 to 26.2) and 37.7 (35.5 to 39.8) against laboratory-confirmed SARS-CoV-2 infection, in the 80 and more years and 65-79 years respectively. For COVID-19 hospitalisation, bivalent VE was estimated as 41.3 (95%CI: 34.5 to 47.5) and 58.5 (95%CI: 51.9 to 64.2), in the 80 and more years and 65-79 years respectively. Considering COVID-related death, bivalent VE was 50.3 (44.6 to 55.3) and 65.1 (95%CI: 59 to 70.4), in the 80 and more years and 65-79 years respectively (Table 2).

**Table 2.** Bivalent vaccine effectiveness against hospitalisation and death in the population aged 65 and more years.

|  | Person.years | Events | VE (%) | VE 95%CI |
| --- | --- | --- | --- | --- |
| **Infection** |  |  |  |  |
| **65 to 79 years** |  |  |  |  |
| Without bivalent vaccine | 417,784.7 | 20,854 | Reference |  |
| With bivalent vaccine | 642,935.6 | 8,985 | 37.7 | (35.5 to 39.8) |
| **≥80 years** |  |  |  |  |
| Without bivalent vaccine | 118,519.5 | 6,348 | Reference |  |
| With bivalent vaccine | 279,340.5 | 9,746 | 23.2 | (20.1 to 26.2) |
| **Hospitalisation** |  |  |  |  |
| **65 to 79 years** |  |  |  |  |
| Without bivalent vaccine | 426,633.6 | 596 | Reference |  |
| With bivalent vaccine | 649,255.6 | 455 | 58.5 | (51.9 to 64.2) |
| **≥80 years** |  |  |  |  |
| Without bivalent vaccine | 120,764.9 | 644 | Reference |  |
| With bivalent vaccine | 282,939.9 | 1,170 | 41.3 | (34.5 to 47.5) |
| **Deaths** |  |  |  |  |
| **65 to 79 years** |  |  |  |  |
| Without bivalent vaccine | 426,836.7 | 412 | Reference |  |
| With bivalent vaccine | 649,399.8 | 378 | 65.1 | (59 to 70.4) |
| **≥80 years** |  |  |  |  |
| Without bivalent vaccine | 120,944.2 | 633 | Reference |  |
| With bivalent vaccine | 283,204.5 | 1,192 | 50.3 | (44.6 to 55.3) |

**Table 3.** Bivalent vaccine effectiveness against hospitalisation and death in the population aged 65 and more years, by time since bivalent vaccine uptake.

|  | Person.years | Events | VE | VE 95%CI |
| --- | --- | --- | --- | --- |
| Infection |  |  |  |  |
| 65 to 79 years |  |  |  |  |
| without bivalent vaccine | 417784.7 | 20854 | Reference |  |
| Time since bivalent vaccination |  |  |  |  |
| 14 to 97 days | 263050 | 4426 | 47.5 | (45.3 to 49.6) |
| 98 to 181 days | 257676.8 | 2735 | 18.4 | (13.3 to 23.1) |
| ≥ 182 days | 122208.8 | 1824 | 0.9 | (-7.1 to 8.3) |
| ≥80 years |  |  |  |  |
| without bivalent vaccine | 118519.5 | 6348 | Reference |  |
| Time since bivalent vaccination |  |  |  |  |
| 14 to 97 days | 106373.2 | 4552 | 28.9 | (25.5 to 32.2) |
| 98 to 181 days | 102143.8 | 3024 | 14.4 | (8.9 to 19.5) |
| ≥ 182 days | 70823.5 | 2170 | 11.3 | (4 to 18) |
| Hospitalisation |  |  |  |  |
| 65 to 79 years |  |  |  |  |
| without bivalent vaccine | 426633.6 | 596 | Reference |  |
| Time since bivalent vaccination |  |  |  |  |
| 14 to 97 days | 265792.9 | 202 | 66.5 | (59.8 to 72.2) |
| 98 to 181 days | 260169.5 | 173 | 46.5 | (32.7 to 57.5) |
| ≥ 182 days | 123293.2 | 80 | 33.8 | (7.1 to 52.9) |
| ≥80 years |  |  |  |  |
| without bivalent vaccine | 120764.9 | 644 | Reference |  |
| Time since bivalent vaccination |  |  |  |  |
| 14 to 97 days | 107332.6 | 518 | 46.9 | (39.3 to 53.6) |
| 98 to 181 days | 103574.4 | 440 | 34.6 | (23.3 to 44.2) |
| ≥ 182 days | 72032.9 | 212 | 29.8 | (11.3 to 44.4) |
| Deaths |  |  |  |  |
| 65 to 79 years |  |  |  |  |
| without bivalent vaccine | 426836.7 | 412 | Reference |  |
| Time since bivalent vaccination |  |  |  |  |
| 14 to 97 days | 265830.9 | 130 | 75.1 | (68.8 to 80) |
| 98 to 181 days | 260231.5 | 169 | 49.4 | (35.4 to 60.3) |
| ≥ 182 days | 123337.4 | 79 | 52.7 | (33.4 to 66.5) |
| ≥80 years |  |  |  |  |
| without bivalent vaccine | 120944.2 | 633 | Reference |  |
| Time since bivalent vaccination |  |  |  |  |
| 14 to 97 days | 107380.8 | 479 | 55.7 | (49.2 to 61.3) |
| 98 to 181 days | 103686.3 | 434 | 45.7 | (36.4 to 53.6) |
| ≥ 182 days | 72137.4 | 279 | 39.3 | (25.1 to 50.8) |

We observed a decline in bivalent VE over time for all outcomes in both age groups. Namely, for 80 or more years, VE against infection decreased 28.9% 14-97 days following vaccination to 11.3% after 182 or more days of vaccination. A similar trend was verified for 65-79 years old, though VE was not significant after 181 days.

Considering COVID-19 hospitalisation, 14-97 days after bivalent vaccine uptake VE reached the maximum of 66.5% and 46.9% for 65-79 and 80 or more years old, respectively, and declined after 182 or more days after vaccination to 33.8% and 29.8%, respectively. Finally, VE against COVID-19-related death peaked at 75.1% for 65-79 years old and decreased to 52.7% after 182 days of vaccine uptake. The corresponding figures for 80 and more years old were 55.7%, 14-79 days after vaccination and 39.3%, 182 and more days after vaccination.

## Discussion

Taking advantage of data collected through electronic health records, this study estimated bivalent vaccine effectiveness in September 2022-May 2023 in a nationwide cohort of Portuguese residents aged 65 or more years old, considering outcomes of different severity levels. Our results indicate that bivalent boosters provide low-to-moderate protection against laboratory-confirmed SARS-CoV-2 infection and moderate protection against severe COVID-19 outcomes.

VE estimates were higher for 65-79 years compared to ≥80 years for all outcomes included in this study. Several factors could contribute to observed differences in levels of conferred protection, such as immunosenescence and deterioration in immune system function due to age. The vaccination campaign rollout in Portugal also resulted in different vaccination histories for these age groups. In response to BA.5 Omicron emergence, national authorities recommended 2^nd^ booster for the ≥80 years population in mid-May 2022, while those 65-79 years old were not targeted by this recommendation. By week 36, when seasonal vaccination with bivalent vaccines started 2^nd^ booster coverage in the ≥80 years group reached 68%, so the reference group in ≥80 years included individuals that were more recently vaccinated compared to 65-79 (who were targeted by a 1^st^ booster in November-December 2021). Thus, the lower VE estimates observed in the ≥ 80 year-olds should also be interpreted in light of the reference group used for the estimation, i.e., individuals with high vaccine coverage (both primary series and multiple boosters). Additionally, vaccines administrated at the beginning of the bivalent vaccine campaign included the BA1/2 component, as ≥80 years were prioritised for vaccination. This age group might have a higher proportion of the BA1/2 vaccine, that mismatched with circulating SARs-CoV-2 variant in early season. In other studies, the BA1/2 vaccine has been shown less effective compared to BA4/5.

We obtained similar results in VE waning compared with previously observed for primary vaccination and booster doses, with levels of protection gradually declining over time (12). This might be due to the natural wane with time and changes in the circulating variant. XXQ became predominant in Portugal by week 10/2023. The decline in VE was in line with other studies (13,14). Compared with these studies, we had a larger follow-up period, providing a more comprehensive assessment of VE waning over time.

Our study has some limitations that should be acknowledged. First, it is based on routine healthcare databases not collected for research purposes. Previous infection was included in the analysis as a confounder. However, asymptomatic infections and self-tests done at home might not be reported, thus underestimating the number of individuals with previous infections. Thus, some individuals without bivalent vaccines could have had a recent infection, reducing VE, as natural immunity would still protect them.

## Data Availability

All data supporting the findings presented were obtained from population registries that belong to the General Directorate for Health (DGS), Central Administration of the Health System (ACSS) and Shared Services of the Ministry of Health (SPMS). The data has sensitive information and was licensed for exclusive use in the current study. Due to privacy regulations, the data is not openly available.

